# Declining Trend of Sudden Cardiac Death in Younger Individuals: A 20–Year Nationwide Study

**DOI:** 10.1101/2024.03.14.24304325

**Authors:** Carl Johann Hansen, Jesper Svane, Peder Emil Warming, Thomas Hadberg Lynge, Rodrigue Garcia, Carolina Malta Hansen, Christian Torp-Pedersen, Jytte Banner, Bo Gregers Winkel, Jacob Tfelt-Hansen

## Abstract

**Background:** Declining all-cause and cardiovascular mortality rates have been well-documented, yet temporal trends of sudden cardiac death (SCD) in the young are unclear. We provide contemporary nationwide estimates of the incidence and temporal trends of SCD in the young aged 1–35 from 2000–2019 and evaluate these trends in relation to changes in out-of-hospital cardiac arrest (OHCA) patterns and implantable cardioverter defibrillator (ICD) implantations.

**Method:** All individuals aged 1–35 living in Denmark from 2000–2019 were included. Adjudication of SCD cases relied on multiple sources, including death certificates, medical files, and autopsy reports. All OHCA were captured in a nationwide prospective registry, and ICD implantations were registered through administrative registries. Crude and sex-and age-standardized annual incidence rates of SCD were calculated, and temporal changes in SCD incidence were computed as percentage change annualized (PCA). Trends in OHCA survival and characteristics, as well as ICD implantations, were assessed.

**Results:** During the 20-year study period (47.5 million person-years), 1 057 SCD were identified (median age 29 years, 69% male sex). The overall incidence of SCD was 2.2 per 100 000 person-years and declined by 3.31% (95%CI 2.42 to 4.20) annually, corresponding to a 49.0% (95%CI 38.7 to 57.6) reduction during the study. Rates of witnessed SCD declined markedly (PCA −7.03% [95%CI −8.57 to −5.48]), but we observed no changes in the rate of unwitnessed SCD (PCA −0.09% [95%CI −1.48 to 1.31]). Consequently, the proportion of unwitnessed SCD increased by 79% (p<0.001). Survival after OHCA in young individuals aged 1–35 increased from 3.9% to 28% in the same time frame, mainly attributable to increased bystander CPR and defibrillation. The implantation rate of ICD increased from 0.76 to 1.55 per 100 000 PY, and the prevalence of young individuals with an ICD increased 16-fold.

**Conclusion:** SCD incidence rates in the young declined by 49% over the last two decades. The decline was paralleled by improved survival in OHCA victims and higher ICD implantation rates. However, rates of unwitnessed SCD were unchanged, which calls for new perspectives in preventive strategies.

**Clinical Perspectives:** *What is new? (max 100 words, 2-3 bullets):*

- Over the past 20 years, the incidence of SCD in young individuals aged 1–35 declined by 49%.
- Concomitantly, the 30-day survival after OHCA increased from 4% to 28%, driven by improved bystander CPR rates reaching +80%, and ICD implantations in young adults increased markedly.
- We observed no decline in the incidence of unwitnessed SCD during the study period.

*What are the clinical implications (max 100 words, 2-3 bullets):*

- As the majority (73%) of SCD in the most recent years were unwitnessed, preventive strategies targeting unwitnessed cardiac arrests need further attention. Incorporating connected devices, e.g. smart watches, in prevention could potentially help identify ventricular arrhythmias and thus improve survival among unwitnessed cardiac arrest.
- Low bystander AED rates (8%) call for further improvement in early AED deployment.

## Introduction

While overall and cardiovascular disease mortality rates in Western countries have declined in recent decades^1,2^, sudden cardiac death (SCD) among young individuals aged ≤35 years continues to pose a significant public health challenge associated with vast psychological consequences and a substantial loss of productive years^3,4^.

Previous studies have shown that the incidence rates (IR) of SCD among young individuals range from 1–3 per 100 000 person-years^5,6^, yet there is limited research on contemporary data, and previous studies on temporal trends have been conflicting^7–11^. As the causes and circumstances of cardiac arrest are strongly age-dependent^12^, temporal trends in the elderly population cannot reliably be extrapolated to the young. Given the decline in mortality rates over the past decades, contemporary estimates of SCD incidence are crucial for evaluating and implementing specific preventive health initiatives targeting the young.

In the current study, we aimed to provide contemporary estimates of the incidence of SCD in the young aged 1–35 years and evaluate the temporal trends from 2000–2019 in relation to changes in out-of-hospital cardiac arrest (OHCA) patterns and implantable cardioverter defibrillator (ICD) implantations. Furthermore, we aimed to identify subgroups of the SCD population, in which current preventive strategies have had less impact and where targeted preventive initiatives could be beneficial.

## Methods

This nationwide observational study complies with the STROBE (Strengthening the Reporting of Observational Studies in Epidemiology) guidelines for observational studies.

### Data availability

Data are owned by a third party, and the authors do not have the right to share data; however, upon request, the authors will help in applying to the third party for approval of data sharing.

### Data sources

All Danish citizens are assigned a unique civil registration number, which allows data linkage at the individual level across the administrative registries in Denmark. These registers are government-maintained and comprise longitudinally collected administrative and health-care data^13^.

Information from the following registries was included: *The Danish Civil Registration System* holds information on sex, migration, and vital status and was used to identify the study population^13^. *The Danish Register of Causes of Death* holds information on the causes and circumstances of all deaths in Denmark^14^. *The National Patient Registry* holds information on all hospital encounters and procedures (in-patient since 1978 and out-patient since 1995) and was used to identify comorbidities and ICD implantations^15^. *The Danish National Prescription Registry* holds information on prescriptions redeemed at Danish pharmacies since 1995 and was used to identify comorbidities and comedication^16^.

Additional data sources were a clinical database containing detailed information on all young SCD aged 1–35 in Denmark from 2000–2019^6^, and the *Danish Cardiac Arrest Registry*, which comprises information on all out-of-hospital cardiac arrests (OHCA) in Denmark since 2001 with attempted resuscitation by emergency medical services^17^.

### Study population and characteristics

All individuals aged 1–35 years living in Denmark during 2000–2019 were included. The annual at-risk population was identified on 1 January of each year (index date).

SCD cases were identified through a multi-source approach, which has previously been described in detail^6,18^. Adjudication was based on manual examination of all death certificates, autopsy reports, and discharge summaries preceding death during the study period. SCD was defined as the *sudden, unexpected death of presumed cardiac origin that occurs within 1 hour of symptom onset in witnessed cases and within 24 hours of last seen alive and well in unwitnessed cases*^19^.

To contextualize the trends in SCD, we examined trends in 1) survival and bystander interventions in all OHCA aged 1–35 registered in the Danish Cardiac Arrest Registry from 2001-2019 and 2) all ICD implantations registered during the study period in individuals aged 1–35. ICD implantations were identified by procedural codes from the National Patient Registry (ICD-10 code: BFCB0) and subsequently categorized as either primary prophylactic (no previous record of a cardiac arrest) or secondary prophylactic (in patients with a previous cardiac arrest).

Comorbidities were identified from the *Danish National Patient Registry* as any hospital contact with a given diagnosis in the ten years preceding the index date; additionally, hypertension, diabetes, and psychiatric disease were identified by recent co-medication^20,21^. *Any comorbidity* was defined as the presence of hypertension, cardiovascular disease (CVD), chronic kidney disease (CKD), diabetes, COPD, epilepsy, and psychiatric disease (see *Suppl. Table 1* for definitions of comorbidity). Information on comorbidities was available until 2018.

### Statistical analyses

The study period was divided into 4-year intervals (2000-2003, 2004-2007, 2008-2011, 2012-2015, and 2016-2019) to compare characteristics over time. For descriptive statistics, categorical variables were presented as count and percentages and continuous variables as median and interquartile range (IQR). Temporal trends in continuous variables were assessed with linear regression and presented as 20-year absolute change, and in categorial variables with logistic regression with year as a covariate and presented as 20-year relative risk.

IR were calculated as the ratio between mortality counts and the appropriate background population with confidence intervals calculated by Poisson’s exact method. Annual IR were standardized on age and sex by direct standardization, using 5-year age groups and the average Danish population throughout the study period as the standard population. Confidence intervals for directly standardized incidence rates (DSIR) were calculated based on the gamma distribution^22^. Temporal trends in DSIR were calculated as the percentage change annualized (PCA) by fitting log-linear models to the DSIR, using weighted least square regression^23^. PCA values were compared by a Wald test of the log-linear models.

A two-sided p-value of 5% was considered significant. All calculations were performed in R v. 4.2.1 (R Foundation for Statistical Computing, Vienna, Austria) using the following packages: *data.table, targets, tidyverse, heaven, and gtsummary*.

### Ethics

This study complies with the Helsinki Declaration and was approved by the Regional Data Committee (P-2019-813 and P-2019-523). Registry-based studies on deidentified data are exempt from ethical approvals in Denmark. Due to data regulations, counts below four have been masked (<=3) throughout the manuscript.

## Results

During the 20-year study period, 4 274 630 unique individuals were observed for 47.5 million person-years (PY). In total, 15 186 deaths occurred, of which 1 057 (7%) were categorized as SCD.

### Trends in population characteristics

The average population in Denmark aged 1–35 was 2.38 mill (median age: 18 years [IQR 10–27], 51% male), and both population size, age, and sex distribution remained stable throughout the study (*Suppl. Table 2*). While the overall comorbidity burden was low, the prevalence of most comorbidities increased annually. Notably, the prevalence of individuals with a previous cardiac arrest or an ICD increased significantly during the study period (estimated 20-year RR: 8.05 (p<0.001) and 11.03 (p<0.001), respectively).

Clinical characteristics of the 1 057 SCD (median age: 29 [IQR 22–33], 69% male) stratified by time period are shown in *Table 1*. Throughout the study period, there was a trend towards a higher proportion of male sex (estimated 20-year RR: 1.13, p = 0.085). The prevalence of CVD remained constant (15%, p for trend = 0.55). The autopsy rate was stable at 67% (p for trend 0.64), and no substantial changes in causes of death were observed. The proportion of unwitnessed SCD increased by 79% during the study period (estimated 20-year RR: 1.79, p<0.001).

**Table 1:**
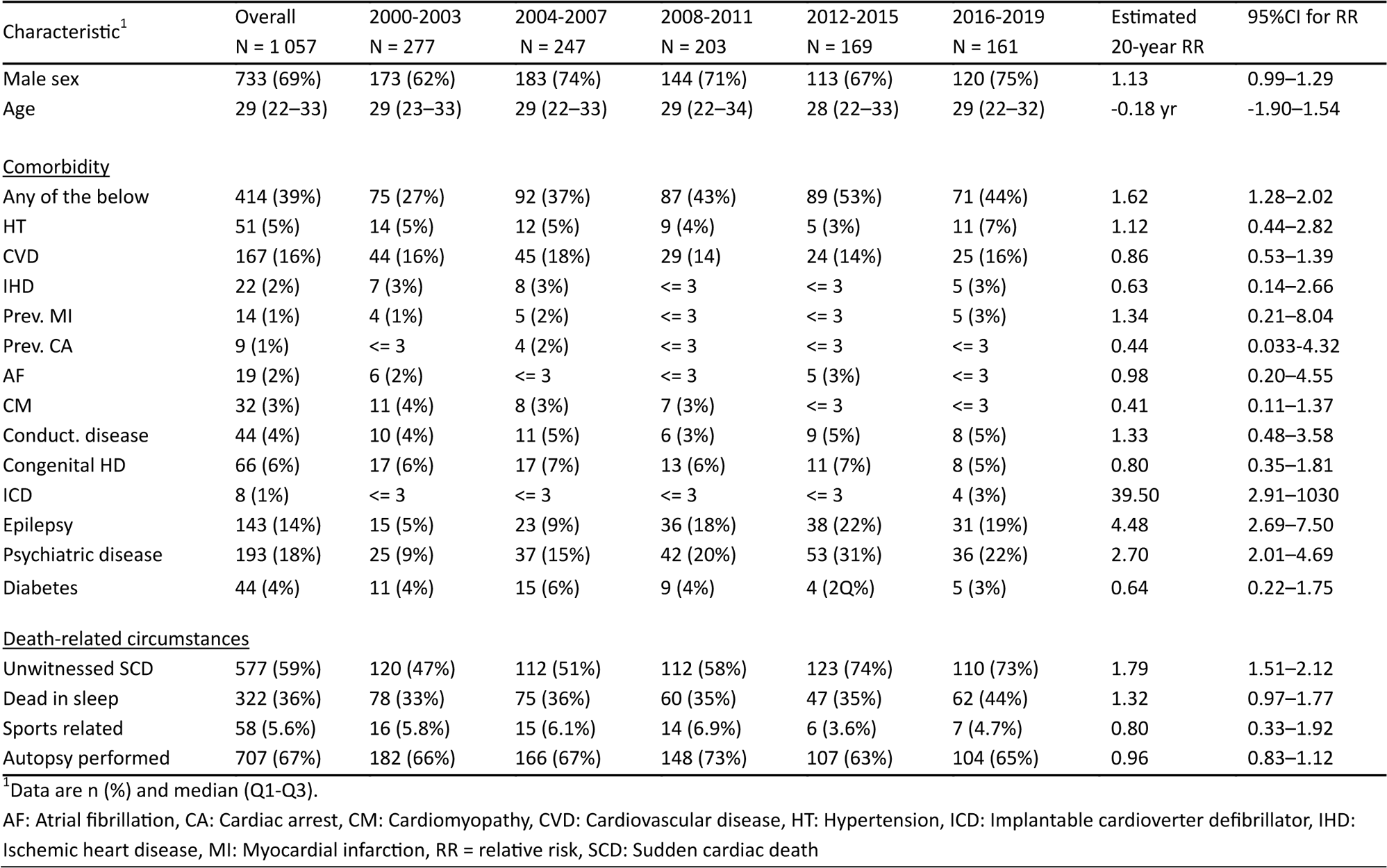
Characteristics of all sudden cardiac death (SCD) cases, stratified by time period.

| Characteristic<sup>1</sup> | Overall<br>N = 1 057 | 2000-2003<br>N = 277 | 2004-2007<br>N = 247 | 2008-2011<br>N = 203 | 2012-2015<br>N = 169 | 2016-2019<br>N = 161 | Estimated<br>20-year RR | 95%CI for RR |
| --- | --- | --- | --- | --- | --- | --- | --- | --- |
| Male sex | 733 (69%) | 173 (62%) | 183 (74%) | 144 (71%) | 113 (67%) | 120 (75%) | 1.13 | 0.99–1.29 |
| Age | 29 (22–33) | 29 (23–33) | 29 (22–33) | 29 (22–34) | 28 (22–33) | 29 (22–32) | -0.18 yr | -1.90–1.54 |
| <u>Comorbidity</u> |  |  |  |  |  |  |  |  |
| Any of the below | 414 (39%) | 75 (27%) | 92 (37%) | 87 (43%) | 89 (53%) | 71 (44%) | 1.62 | 1.28–2.02 |
| HT | 51 (5%) | 14 (5%) | 12 (5%) | 9 (4%) | 5 (3%) | 11 (7%) | 1.12 | 0.44–2.82 |
| CVD | 167 (16%) | 44 (16%) | 45 (18%) | 29 (14) | 24 (14%) | 25 (16%) | 0.86 | 0.53–1.39 |
| IHD | 22 (2%) | 7 (3%) | 8 (3%) | <= 3 | <= 3 | 5 (3%) | 0.63 | 0.14–2.66 |
| Prev. MI | 14 (1%) | 4 (1%) | 5 (2%) | <= 3 | <= 3 | 5 (3%) | 1.34 | 0.21–8.04 |
| Prev. CA | 9 (1%) | <= 3 | 4 (2%) | <= 3 | <= 3 | <= 3 | 0.44 | 0.033–4.32 |
| AF | 19 (2%) | 6 (2%) | <= 3 | <= 3 | 5 (3%) | <= 3 | 0.98 | 0.20–4.55 |
| CM | 32 (3%) | 11 (4%) | 8 (3%) | 7 (3%) | <= 3 | <= 3 | 0.41 | 0.11–1.37 |
| Conduct. disease | 44 (4%) | 10 (4%) | 11 (5%) | 6 (3%) | 9 (5%) | 8 (5%) | 1.33 | 0.48–3.58 |
| Congenital HD | 66 (6%) | 17 (6%) | 17 (7%) | 13 (6%) | 11 (7%) | 8 (5%) | 0.80 | 0.35–1.81 |
| ICD | 8 (1%) | <= 3 | <= 3 | <= 3 | <= 3 | 4 (3%) | 39.50 | 2.91–1030 |
| Epilepsy | 143 (14%) | 15 (5%) | 23 (9%) | 36 (18%) | 38 (22%) | 31 (19%) | 4.48 | 2.69–7.50 |
| Psychiatric disease | 193 (18%) | 25 (9%) | 37 (15%) | 42 (20%) | 53 (31%) | 36 (22%) | 2.70 | 2.01–4.69 |
| Diabetes | 44 (4%) | 11 (4%) | 15 (6%) | 9 (4%) | 4 (2Q%) | 5 (3%) | 0.64 | 0.22–1.75 |
| <u>Death-related circumstances</u> |  |  |  |  |  |  |  |  |
| Unwitnessed SCD | 577 (59%) | 120 (47%) | 112 (51%) | 112 (58%) | 123 (74%) | 110 (73%) | 1.79 | 1.51–2.12 |
| Dead in sleep | 322 (36%) | 78 (33%) | 75 (36%) | 60 (35%) | 47 (35%) | 62 (44%) | 1.32 | 0.97–1.77 |
| Sports related | 58 (5.6%) | 16 (5.8%) | 15 (6.1%) | 14 (6.9%) | 6 (3.6%) | 7 (4.7%) | 0.80 | 0.33–1.92 |
| Autopsy performed | 707 (67%) | 182 (66%) | 166 (67%) | 148 (73%) | 107 (63%) | 104 (65%) | 0.96 | 0.83–1.12 |

<sup>1</sup>Data are n (%) and median (Q1-Q3).
AF: Atrial fibrillation, CA: Cardiac arrest, CM: Cardiomyopathy, CVD: Cardiovascular disease, HT: Hypertension, ICD: Implantable cardioverter defibrillator, IHD: Ischemic heart disease, MI: Myocardial infarction, RR = relative risk, SCD: Sudden cardiac death

### SCD Incidence rates and temporal trends

SCD Incidence rates are presented in *Table 2.* The overall SCD IR throughout the entire study period was 2.2 per 100 000 PY (95%CI 2.1-2.4). Crude SCD rates decreased from 2.89 per 100 000 PY (95%CI 2.56–3.24) in 2000–2003 to 1.67 (95%CI 1.42–1.94) per 100 000 PY in 2016–2019. After sex- and age adjustment, the annual decline (percentage change annualized [PCA]) was −3.31% (95%CI −4.20; −2.42), accumulating to a 49.0% (95%CI 38.7–57.6) reduction in the SCD incidence over the 20-year period (*Figure 1*).

**Figure 1:**
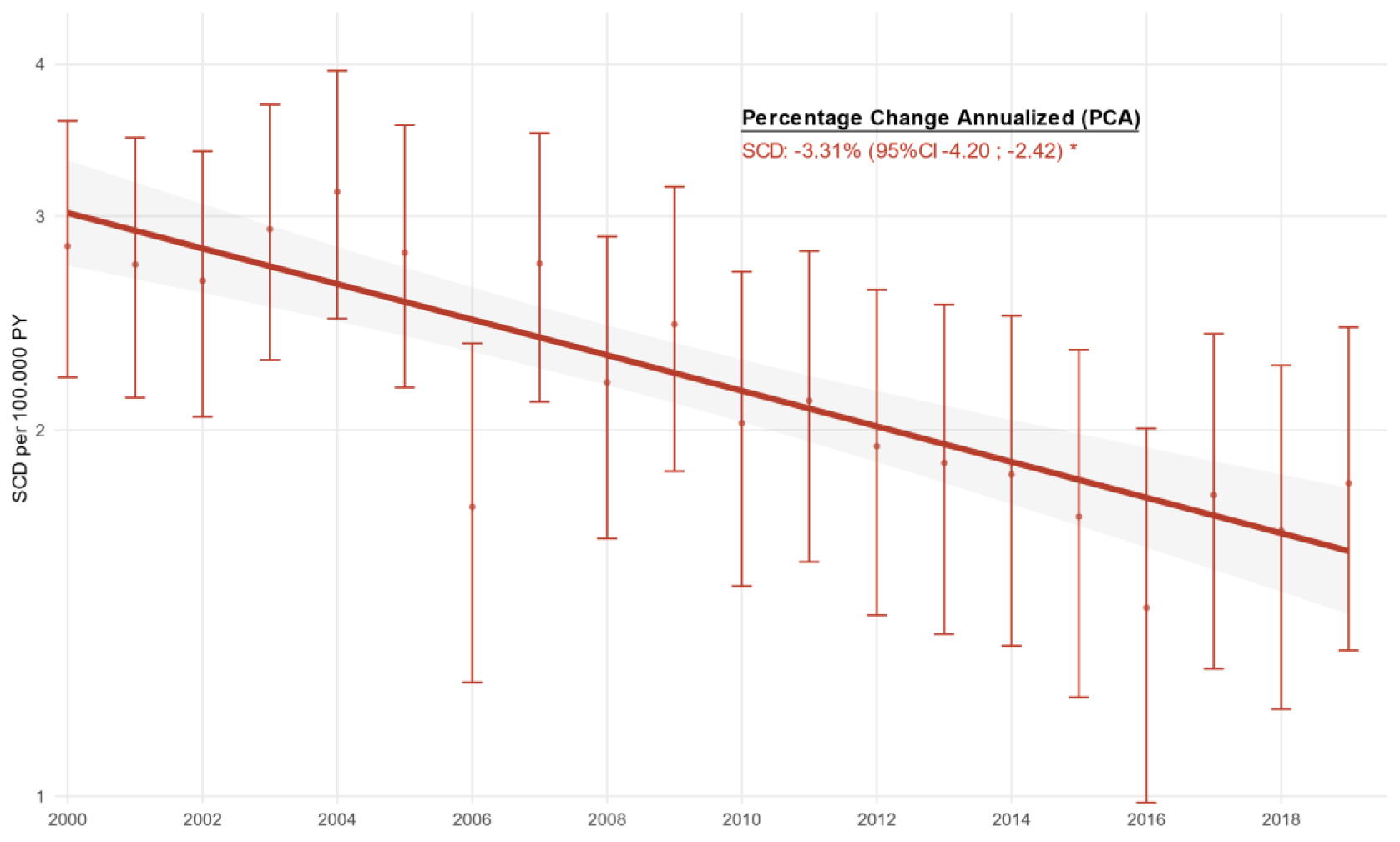
Temporal trends in sudden cardiac death (SCD) incidence from 2000–2019

**Table 2:** Incidence rates and temporal trends of sudden cardiac death (SCD) in the young aged 1–35.

|  | SCD cases | Person-years | Crude incidence estimates |  |  |  | Adjusted annual changes |  |
| --- | --- | --- | --- | --- | --- | --- | --- | --- |
|  |  |  | Crude IR | IR 95%CI | IRR | IRR 95%CI | PCA (%) | PCA 95%CI |
| All SCD | 1 057 | 47.5 million | 2.2 | 2.1-2.4 | - | - | -3.31 | -4.2 to -2.42 |
| By groups |  |  |  |  |  |  |  |  |
| Time period |  |  |  |  |  |  |  |  |
| 2000-2003 | 277 | 9.6 million | 2.89 | 2.56-3.24 | - | - | - | - |
| 2004-2007 | 247 | 9.4 million | 2.6 | 2.3-3.0 | 0.91 | 0.77-1.1 | - | - |
| 2008-2011 | 203 | 9.4 million | 2.2 | 1.9-2.5 | 0.75 | 0.62-0.89 | - | - |
| 2012-2015 | 169 | 9.4 million | 1.8 | 1.5-2.1 | 0.62 | 0.51-0.75 | - | - |
| 2016-2019 | 161 | 9.7 million | 1.67 | 1.42-1.94 | 0.58 | 0.47-0.7 | - | - |
| Sex |  |  |  |  |  |  |  |  |
| Female | 324 | 23.3 million | 1.4 | 1.2-1.5 | - | - | -4.66 | -6.17 to -3.14 |
| Male | 733 | 24.2 million | 3.0 | 2.8-3.3 | 2.2 | 1.9-2.5 | -2.73 | -3.95 to -1.50 |
| Age |  |  |  |  |  |  |  |  |
| 1-18 | 175 | 23.8 million | 0.74 | 0.63-0.85 | - | - | -4.7 | -7.5 to -1.9 |
| 19-35 | 882 | 23.7 million | 3.70 | 3.50-4.00 | 5.1 | 4.3-6.0 | -2.8 | -3.9 to -1.8 |
| Witnessed status |  |  |  |  |  |  |  |  |
| Witnessed | 406 | 47.5 million | 0.85 |  | - | - | -7.03 | -5.46 to -2.71 |
| Unwitnessed | 651 | 47.5 million | 1.37 |  | 1.6 | - | -0.09 | -1.48 to 1.31 |
| CI: Confidence interval, IR: Incidence rates, IRR: Incidence rate ratio, PCA: Percentage change annualized, SCD: Sudden cardiac death |  |  |  |  |  |  |  |  |

We observed marked differences in SCD incidence according to sex and witnessed status. Males were at considerably higher risk of SCD (IRR: 2.2 [95%CI 1.9–2.5, p<0.001]), and the was a trend towards an increasing discrepancy between the sexes throughout the study period, illustrated by a 1.9% lower PCA in males compared to females (p for difference = 0.052) (*Figure 2A*). The incidence of witnessed SCD decreased significantly with a PCA of −7.03% (95%CI −8.57; −5.48). Contrarily, we observed no difference in the incidence of unwitnessed SCD (PCA −0.09% [95%CI – 1.48;1.31])(*Figure 2B*).

**Figure 2:**
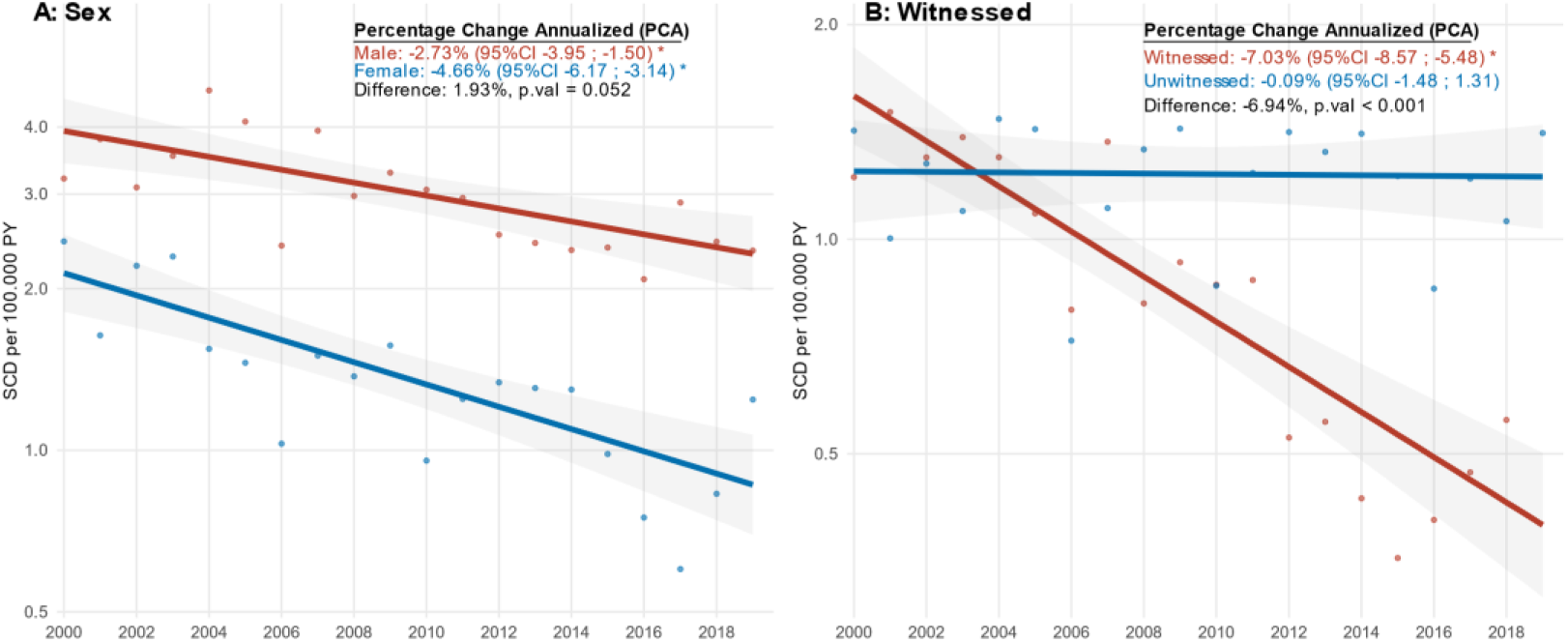
Temporal trends in sudden cardiac death (SCD) incidence, by sex (panel A) and witnessed status (panel B).

### Out-of-hospital cardiac arrest

From 2001–2019, 2 256 OHCA in individuals aged 1–35 years were registered (median age 25 [IQR 19–31], 70% male sex). Comparing the first and the last time periods, we observed increasing proportions of witnessed OHCA (28% to 39%, p<0.001), bystander cardiopulmonary resuscitation (CPR) (30% to 79%, p<0.001), bystander defibrillation (1.8% to 7.7%, p<0.001) and 30-day survival (3.9% to 28%, p<0.001) (*Figure 3*). Overall, OHCA characteristics were similar between both sexes except for females being younger (24 vs 26 years, p = 0.014) and OHCA in women more often occurring in private residences (67% vs 56%, p<0.001). 30-day survival was significantly better among females (22% vs 16%, p <0.001). When temporal changes in OHCA characteristics were assessed, bystander CPR and defibrillation rates improved markedly more among women and surpassed the rates among males (*Suppl Table 3*).

**Figure 3:**
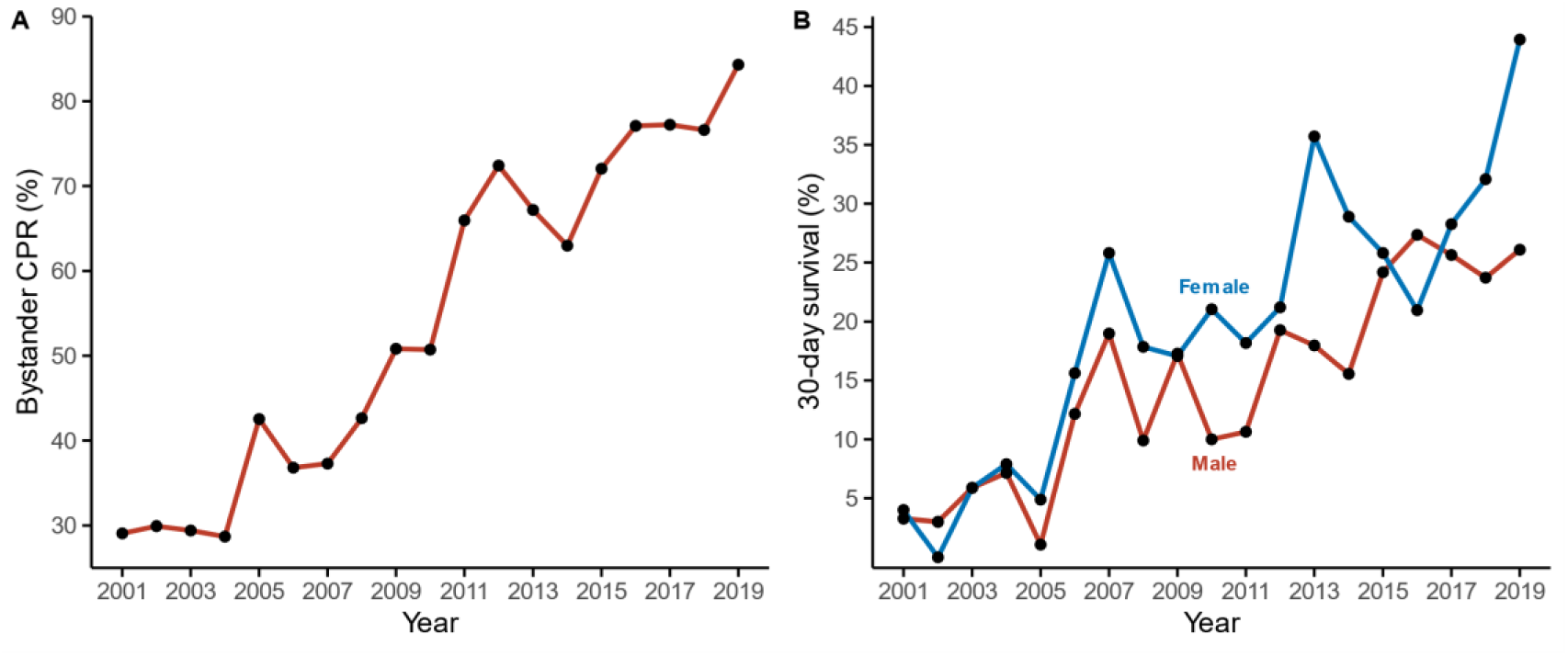
Temporal trends in bystander cardiopulmonary resuscitation (CPR) (Panel A) and cardiac arrest survival (panel B) in the young aged 1–35.

### ICD implantation

During the 20-year study period, 606 ICDs (406 primary prevention, 200 secondary prevention) were implanted in individuals aged 1–35 years (*Figure 4*). The ICD-implantation rate doubled from 73 implantations in 2000-2003 (IR 0.76 per 100 000 PY) to 150 in 2016-2019 (IR 1.55 per 100 000 PY), and the proportion of secondary preventive ICDs increased from 19% to 38%.

**Figure 4:**
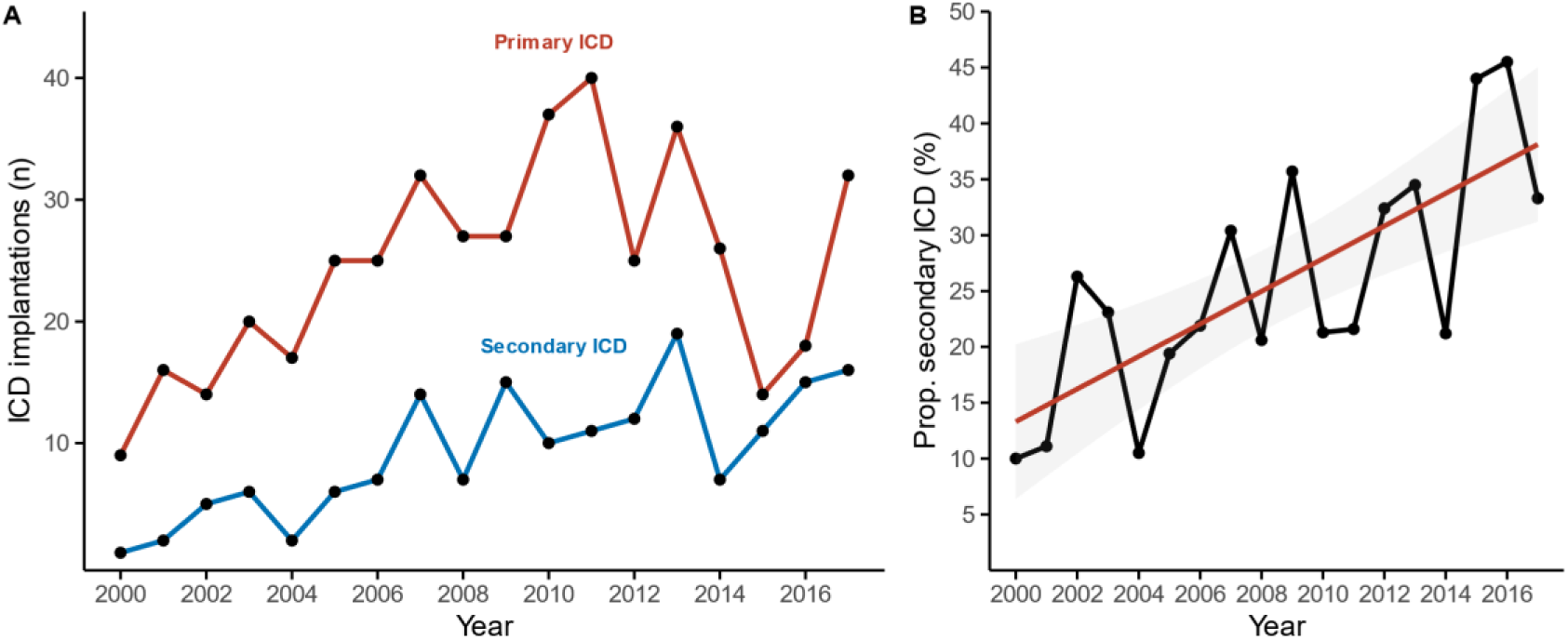
Temporal trends in implantable cardioverter defibrillator implantation in the young aged 1–35. Panel A: Implantations of primary and secondary preventive ICD. Panel B: Trends in the proportion of secondary preventive ICD of all ICDs implanted.

## Discussion

This longitudinal, study provides contemporary nationwide incidence estimates for SCD based on a large cohort of young SCD cases aged 1–35 years in Denmark. The study’s principal findings include a significant decrease in the incidence of SCD (49% over the last 20 years), a substantially 7-fold improved survival following OHCA in young individuals and higher rates of ICD implantation. We believe this is among the first studies to contextualize the trends in SCD to OHCA survival and ICD implantations in young individuals. Notably, the study provides an up-to-date overview of the epidemiology of SCD with potential implications for future preventive initiatives to reduce SCD.

### Incidence rates of SCD

The reported IR of 1.67 per 100 000 PY in the most recent time period (2016–2019) is comparable to the IR reported in the literature^5,10,24,25^. Direct cross-study comparison of incidence rates is difficult due to vast differences in methodology and study populations. Our multi-source identification of SCD, based on all deaths in Denmark from 2000–2019, reduces the risk of selection bias typically associated with epidemiological studies on selected populations (autopsy cohorts) or ICD-codes from death certificates, and we believe it offers the most accurate estimate of the true burden of SCD.

More importantly, we observed a 3.3% annual decline in SCD incidence, accumulating to a 49% reduction in SCD over the 20-year study period. All observed subgroups exhibited declining rates, apart from unwitnessed SCD, where the IR remained stable. Our group has previously reported temporal trends in young SCD from 2000-2009^9^, and the trend has continued to decline at a comparable rate over the past decade. Similar findings have been reported in young SCD^7,26^, SCD in all ages^2,11^, and cardiovascular mortality^2^, supporting an overall trend of declining SCD mortality.

### Improved survival after OHCA

The reduction in SCD mortality is multifactorial, but improved survival after OHCA is among the most important contributors. Several strategies to improve OHCA survival have been implemented in Denmark in the last decades, including widespread CPR training, lay-person activation (“Heart runner” program), and increased public automated external defibrillators (AED) availability^27,28^, and the effects of these community-based interventions on survival are well documented^29,30^. During the study period, survival after OHCA in individuals aged 1–35 increased from 4% to 28%, concomitantly with an increase in bystander CPR and defibrillation to approximately 80% and 8%, respectively. Although a significant proportion of OHCA in young individuals are non-cardiac^31^, this improved survival translates directly into a lower SCD incidence. Although the rate of bystander defibrillation increased significantly during the study period, AEDs were only used in 8% of the young OHCA victims. The low rate was likely due to most arrests occurring in private homes, where rates of bystander defibrillation remain low^17^. Early bystander defibrillation is a key prognostic factor for survival^29^, and further improvement in the rate of bystander AED use is critical in improving survival.

The proportion of female SCD decreased during the study period, although just missing statistical significance (p = 0.058) – we think this is likely an issue of statistical power. Similar findings have been described in studies of both younger^10^ and older^11^ SCD cohorts, and higher OHCA survival rates have been reported in females^32,33^. In the current study, bystander CPR and defibrillation rates — key prognostic markers for improved survival — increased among both sexes, but the increase was markedly higher among women, where the rates even surpassed those among males. This difference likely contributed to the observed discrepancy between sexes in OHCA survival and SCD incidence. The equal CPR and defibrillation rates in recent years contrast previous reports of OHCA in females^33,34^ and could reflect differences in study populations and characteristics (younger age in the current study) or cultural differences towards resuscitation attempts in younger women.

### Prevention of SCD and identification of high-risk individuals

ICD implantation is a key component in preventing SCD in high-risk patients, as it significantly reduces the risk of arrhythmic death^19,35^. During the study period, the prevalence of ICDs in the young general population increased more than 10-fold, which is likely multifactorial. First, as the number of OHCA survivors increases, more patients will be eligible for ICD implantation. Causes of OHCA among young individuals differ from the elderly with more structural and inherited cardiac disease^12,18^, in whom secondary ICD implantation is more likely. Accordingly, we observed an increased ICD implantation in cardiac arrest survivors. Similar findings have been reported in Western Australia, where ICD implantation rates increased 15-fold from 1995 to 2009^11^, paralleled by improved OHCA survival. Second, substantial improvements in our understanding of the pathophysiology and genetics underlying inherited cardiac diseases have improved risk stratification significantly. Combined with systematic family cascade screening of relatives to SCA/SCD victims, in whom the risk of inherited cardiac diseases is significantly higher^36^, we likely identify more individuals at risk of SCD who are eligible for a primary preventive ICD.

For both primary and secondary prevention ICDs, whether mortality reduction is due to the actual ICD, improved medical care and awareness in patients with an ICD (performance bias), or a combination remains speculative. However, a recent study by Ruwald et al.^28^ found a significant decline in the delivery of appropriate shocks in patients with secondary ICDs during the last decade, and thus fewer who seem to benefit from their ICD. As young individuals with an ICD have a high lifetime risk of device-related complications^37^, this finding highlights the need for better pre-implantation risk stratification and identification of those who may benefit from an ICD, in order to reduce unnecessary implantations.

A surprising finding of this study was the discrepancy in SCD trends between witnessed and unwitnessed SCD. As we only observed declining rates of SCD that were witnessed, the proportion of unwitnessed SCD increased by 79% to approximately three in four, which is markedly higher than previously reported^38,39^. Previous studies have identified key differences between witnessed and unwitnessed SCD — e.g. more unexplained deaths, higher proportion of males, different comorbidity profiles and more frequent death during sleep among unwitnessed SCD^38,40^. Therefore, this shift has substantial implications for the epidemiology of SCD, and future preventive measures must reflect this change as strategies targeting OHCA may have a limited impact on preventing unwitnessed SCD. Prevention of SCD that occur unwitnessed or during sleep is particularly difficult, as timely recognition and intervention is hindered and survival chances thus dramatically lower. However, connected devices —e.g., smart watches and speakers— are increasingly capable of identifying malignant arrhythmias and alerting first responders or family members^41,42^. Although these solutions are still in their infancy, they present an exciting opportunity to monitor high-risk patients and lower mortality^43,44^.

### Study limitations

We acknowledge several limitations of our study. First, the observational nature imposes limitations on the assertions of causality, and interpretation should be made with caution. Second, changes in coding practices, diagnostic criteria, and procedural indications during the 20-year study period may have impacted the observed trends. However, this would primarily affect estimates of comorbidity and have less impact on SCD rates due to the multi-source identification of SCD. Similarly, the OHCA registry has undergone administrative changes during the study period with improved capture due to electronic registration since 2016. However, the improving trend in survival was observed throughout the entire study period, indicating that the underreporting of OHCA cases was likely at random with little effect on our estimates. We had no information about ICD therapies, which could have added further insight into the value if increased ICD implantation. Last, information on modifiable risk factors (BMI, smoking, alcohol intake, activity level) that may influence disease patterns was not available from the national registries.

## Conclusion

Over the past 20 years, there has been a 49% decrease in the incidence of SCD in young individuals aged 1–35 in Denmark. Steep inclines in both ICD implantations and survival rates of cardiac arrest victims parallelled the decline. However, we observed no change in the incidence of unwitnessed SCD, and preventive efforts need to address this issue, e.g., by concentrating on near-term prevention.

## Non-standard Abbreviations and Acronyms

CA: Cardiac arrest
ICD: Implantable cardioverter-defibrillator
IR: Incidence Rate
IRR: Incidence Rate Ratio
OHCA: Out-of-hospital cardiac arrest
SCD: Sudden Cardiac Death

## Acknowledgements

The authors acknowledge the Danish Clinical Quality Program – National Clinical Registries (RKKP). for making it possible to work with data from the Danish Cardiac Arrest Registry.

## Supplemental Material

**Suppl. Table 1:**
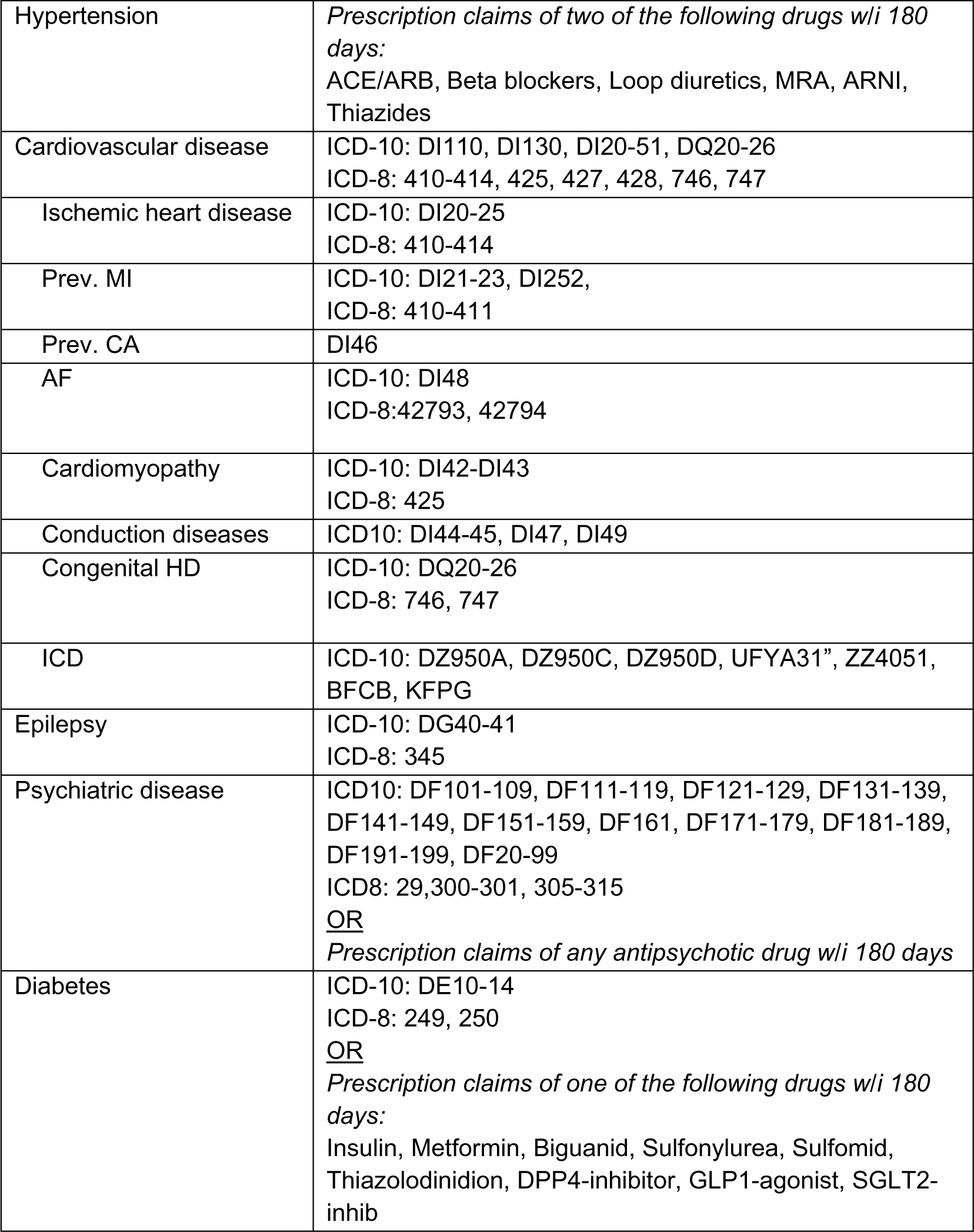
Definitions of comorbidities and ICD-codes used to define diseases.

**Suppl. Table 2:** Characteristics of the general population, stratified by year.

|  | Overall | 2000 | 2001 | 2002 | 2003 | 2004 | 2005 | 2006 | 2007 | 2008 | 2009 |
| --- | --- | --- | --- | --- | --- | --- | --- | --- | --- | --- | --- |
| Population | 47 512 622 | 2 414 649 | 2 403 520 | 2 392 209 | 2 375 476 | 2 360 593 | 2 351 574 | 2 347 356 | 2 347 663 | 2 350 387 | 2 354 962 |
| Male sex | 24 212 851<br>(51%) | 1 233 010<br>(51%) | 1 226 108<br>(51%) | 1 219 309<br>(51%) | 1 209 652<br>(51%) | 1 201 661<br>(51%) | 1 196 358<br>(51%) | 1 193 712<br>(51%) | 1 193 723<br>(51%) | 1 195 151<br>(51%) | 1 197 905<br>(51%) |
| Age | 18 (10–27) | 20 (9–28) | 19 (9–28) | 19 (9–28) | 19 (9–28) | 18 (9–28) | 18 (9–28) | 18 (9–28) | 18 (9–28) | 18 (9–27) | 18 (9–27) |
| <u>Comorbidity</u> |  |  |  |  |  |  |  |  |  |  |  |
| Any | 2 661 419<br>(6%) | 98 914<br>(4%) | 100 907<br>(4%) | 103 599<br>(4%) | 105 834<br>(5%) | 108 175<br>(5%) | 110 685<br>(5%) | 113 408<br>(5%) | 118 171<br>(5%) | 122 296<br>(5%) | 126 577<br>(5%) |
| Hypertension | 57 073<br>(0.1%) | 2 156<br>(<0.1%) | 2 258<br>(<0.1%) | 2 405<br>(0.1%) | 2 485<br>(0.1%) | 2 768<br>(0.1%) | 3 096<br>(0.1%) | 3 195<br>(0.1%) | 3 492<br>(0.1%) | 3 706<br>(0.2%) | 3 811<br>(0.2%) |
| CVD | 542 145<br>(1.1%) | 20 212<br>(0.8%) | 20 979<br>(0.9%) | 21 923<br>(0.9%) | 22 526<br>(0.9%) | 23 011<br>(1.0%) | 23 425<br>(1.0%) | 23 806<br>(1.0%) | 24 481<br>(1.0%) | 25 152<br>(1.1%) | 25 993<br>(1.1%) |
| IHD | 34 359<br>(<0.1%) | 1 393<br>(<0.1%) | 1 487<br>(<0.1%) | 1 633<br>(<0.1%) | 1 670<br>(<0.1%) | 1 750<br>(<0.1%) | 1 829<br>(<0.1%) | 1 822<br>(<0.1%) | 1 832<br>(<0.1%) | 1 809<br>(<0.1%) | 1 800<br>(<0.1%) |
| Prev. MI | 8 464<br>(<0.1%) | 436<br>(<0.1%) | 435<br>(<0.1%) | 459<br>(<0.1%) | 443<br>(<0.1%) | 468<br>(<0.1%) | 468<br>(<0.1%) | 453<br>(<0.1%) | 458<br>(<0.1%) | 457<br>(<0.1%) | 456<br>(<0.1%) |
| Prev. CA | 3 248<br>(<0.1%) | 59<br>(<0.1%) | 58<br>(<0.1%) | 60<br>(<0.1%) | 62<br>(<0.1%) | 74<br>(<0.1%) | 79<br>(<0.1%) | 83<br>(<0.1%) | 101<br>(<0.1%) | 109<br>(<0.1%) | 115<br>(<0.1%) |
| AF | 24 152<br>(<0.1%) | 801<br>(<0.1%) | 891<br>(<0.1%) | 971<br>(<0.1%) | 1 058<br>(<0.1%) | 1 093<br>(<0.1%) | 1 117<br>(<0.1%) | 1 156<br>(<0.1%) | 1 210<br>(<0.1%) | 1 239<br>(<0.1%) | 1 269<br>(<0.1%) |
| CM | 15 285<br>(<0.1%) | 464<br>(<0.1%) | 516<br>(<0.1%) | 560<br>(<0.1%) | 599<br>(<0.1%) | 629<br>(<0.1%) | 667<br>(<0.1%) | 687<br>(<0.1%) | 708<br>(<0.1%) | 741<br>(<0.1%) | 746<br>(<0.1%) |
| Cond. def. | 157 218<br>(0.3%) | 4 251<br>(0.2%) | 4 772<br>(0.2%) | 5 255<br>(0.2%) | 5 727<br>(0.2%) | 6 181<br>(0.3%) | 6 473<br>(0.3%) | 6 746<br>(0.3%) | 7 034<br>(0.3%) | 7 338<br>(0.3%) | 7 620<br>(0.3%) |
| Congenital | 263 384<br>(0.6%) | 11 102<br>(0.5%) | 11 138<br>(0.5%) | 11 331<br>(0.5%) | 11 373<br>(0.5%) | 11 308<br>(0.5%) | 11 348<br>(0.5%) | 11 348<br>(0.5%) | 11 664<br>(0.5%) | 11 995<br>(0.5%) | 12 553<br>(0.5%) |
| ICD | 5 234<br>(<0.1%) | 34<br>(<0.1%) | 47<br>(<0.1%) | 62<br>(<0.1%) | 74<br>(<0.1%) | 105<br>(<0.1%) | 126<br>(<0.1%) | 137<br>(<0.1%) | 152<br>(<0.1%) | 182<br>(<0.1%) | 203<br>(<0.1%) |
| Epilepsy | 422 613<br>(0.9%) | 22 394<br>(0.9%) | 22 498<br>(0.9%) | 22 483<br>(0.9%) | 22 306<br>(0.9%) | 21 807<br>(0.9%) | 21 622<br>(0.9%) | 21 590<br>(0.9%) | 21 528<br>(0.9%) | 21 335<br>(0.9%) | 21 206<br>(0.9%) |
| Psychiatric | 1 291 418<br>(2.7%) | 35 763<br>(1.5%) | 37 237<br>(1.5%) | 39 112<br>(1.6%) | 41 240<br>(1.7%) | 43 576<br>(1.8%) | 45 845<br>(1.9%) | 47 594<br>(2.0%) | 51 258<br>(2.2%) | 54 983<br>(2.3%) | 58 932<br>(2.5%) |
| Diabetes | 262 910<br>(0.5%) | 9 311<br>(0.5%) | 9 419<br>(0.5%) | 9 552<br>(0.5%) | 9 761<br>(0.5%) | 10 204<br>(0.5%) | 10 917<br>(0.5%) | 12 078<br>(0.5%) | 12 938<br>(0.5%) | 13 575<br>(0.5%) | 13 876<br>(0.5%) |
| Deaths | 15 189 | 1 080 | 1 048 | 1 088 | 979 | 952 | 852 | 828 | 843 | 848 | 789 |
| SCD | 1 057 | 71 | 69 | 66 | 71 | 75 | 67 | 41 | 64 | 51 | 57 |

|  | 2010 | 2011 | 2012 | 2013 | 2014 | 2015 | 2016 | 2017 | 2018 | 2019 | Est.<br>20-year<br>RR | 95% CI<br>RR |
| --- | --- | --- | --- | --- | --- | --- | --- | --- | --- | --- | --- | --- |
| Population | 2 356 252 | 2 357 053 | 2 355 283 | 2 356 580 | 2 362 855 | 2 369 955 | 2 392 219 | 2 409 309 | 2 425 936 | 2 428 791 | - | - |
| Male sex | 1 198 188<br>(51%) | 1 198 897<br>(51%) | 1 198 593<br>(51%) | 1 199 630<br>(51%) | 1 204 002<br>(51%) | 1 209 250<br>(51%) | 1 222 689<br>(51%) | 1 232 163<br>(51%) | 1 240 838<br>(51%) | 1 242 012<br>(51%) | 1.003 | 1.003-1.004 |
| Age | 18 (9–27) | 18 (9–27) | 18 (10–27) | 18 (10–26) | 18 (10–26) | 19 (10–26) | 19 (10–27) | 19 (10–27) | 19 (10–27) | 19 (10–27) |  |  |
| <u>Comorbidity</u> |  |  |  |  |  |  |  |  |  |  |  |  |
| Any | 131 658<br>(5.5%) | 137 752<br>(6%) | 143 513<br>(6%) | 149 705<br>(6.5%) | 155 353<br>(6.5%) | 160 769<br>(7%) | 166 271<br>(7%) | 171 486<br>(7%) | 176 917<br>(7.5%) | - | 1.87 | 1.87-1.88 |
| Hypertensio | 3 539<br>(0.2%) | 3 536<br>(0.2%) | 3 356<br>(0.1%) | 3 252<br>(0.1%) | 3 015<br>(0.1%) | 2 763<br>(0.1%) | 2 805<br>(0.1%) | 2 732<br>(0.1%) | 2 703<br>(0.1%) | - | 0.86 | 0.84-0.89 |
| CVD | 26 795<br>(1.1%) | 27 892<br>(1.2%) | 28 911<br>(1.2%) | 29 919<br>(1.3%) | 30 760<br>(1.3%) | 31 783<br>(1.3%) | 33 053<br>(1.4%) | 34 132<br>(1.4%) | 35 131<br>(1.4%) | - | 1.75 | 1.73-1.76 |
| IHD | 1 747<br>(<0.1%) | 1 887<br>(<0.1%) | 1 876<br>(<0.1%) | 1 850<br>(<0.1%) | 1 799<br>(<0.1%) | 1 811<br>(<0.1%) | 1 771<br>(<0.1%) | 1 683<br>(<0.1%) | 1 591<br>(<0.1%) | - | 1.02 | 0.98-1.06 |
| Prev. MI | 430<br>(<0.1%) | 438<br>(<0.1%) | 427<br>(<0.1%) | 409<br>(<0.1%) | 378<br>(<0.1%) | 377<br>(<0.1%) | 391<br>(<0.1%) | 386<br>(<0.1%) | 389<br>(<0.1%) | - | 0.77 | 0.71-0.82 |
| Prev. CA | 135<br>(<0.1%) | 147<br>(<0.1%) | 180<br>(<0.1%) | 214<br>(<0.1%) | 254<br>(<0.1%) | 274<br>(<0.1%) | 292<br>(<0.1%) | 311<br>(<0.1%) | 329<br>(<0.1%) | - | 8.05 | 7.06-9.19 |
| AF | 1 291<br>(<0.1%) | 1 288<br>(<0.1%) | 1 307<br>(<0.1%) | 1 359<br>(<0.1%) | 1 356<br>(<0.1%) | 1 400<br>(<0.1%) | 1 389<br>(<0.1%) | 1 379<br>(<0.1%) | 1 387<br>(<0.1%) | - | 1.50 | 1.44-1.57 |
| CM | 776<br>(<0.1%) | 815<br>(<0.1%) | 843<br>(<0.1%) | 894<br>(<0.1%) | 935<br>(<0.1%) | 934<br>(<0.1%) | 939<br>(<0.1%) | 968<br>(<0.1%) | 987<br>(<0.1%) | - | 1.95 | 1.85-2.06 |
| Cond. def. | 7 922<br>(0.3%) | 8 232<br>(0.3%) | 8 665<br>(0.4%) | 9 117<br>(0.4%) | 9 498<br>(0.4%) | 9 899<br>(0.4%) | 10 395<br>(0.4%) | 10 832<br>(0.4%) | 11 124<br>(0.5%) | - | 2.39 | 2.35-2.44 |
| Congenital | 13 036<br>(0.6%) | 13 568<br>(0.6%) | 14 012<br>(0.6%) | 14 358<br>(0.6%) | 14 688<br>(0.6%) | 15 056<br>(0.6%) | 15 532<br>(0.6%) | 15 982<br>(0.7%) | 16 543<br>(0.7%) | - | 1.58 | 1.55-1.60 |
| ICD | 236<br>(<0.1%) | 268<br>(<0.1%) | 293<br>(<0.1%) | 336<br>(<0.1%) | 398<br>(<0.1%) | 439<br>(<0.1%) | 476<br>(<0.1%) | 551<br>(<0.1%) | 571<br>(<0.1%) | - | 11.03 | 9.92-12.27 |
| Epilepsy | 20 812<br>(0.9%) | 20 656<br>(0.9%) | 20 656<br>(0.9%) | 20 697<br>(0.9%) | 20 725<br>(0.9%) | 20 541<br>(0.9%) | 20 503<br>(0.9%) | 20 304<br>(0.8%) | 20 307<br>(0.8%) | - | 0.86 | 0.85-0.87 |
| Psychiatric | 64 161<br>(2.7%) | 69 324<br>(2.9%) | 74 441<br>(3.2%) | 79 703<br>(3.4%) | 84 552<br>(3.6%) | 88 781<br>(3.7%) | 92 548<br>(3.9%) | 95 602<br>(4.0%) | 98 548<br>(4.1%) | - | 3.06 | 3.04-3.08 |
| Diabetes | 14 330<br>(0.5%) | 14 707<br>(0.5%) | 14 935<br>(0.5%) | 15 105<br>(0.5%) | 15 251<br>(0.5%) | 15 148<br>(0.5%) | 15 261<br>(0.5%) | 15 692<br>(0.5%) | 16 150<br>(0.5%) | - | 1.77 | 1.75-1.79 |
| Deaths | 675 | 685 | 584 | 564 | 590 | 538 | 605 | 560 | 571 | 507 | - | - |
| SCD | 47 | 48 | 44 | 43 | 43 | 39 | 34 | 42 | 40 | 45 | - | - |

**Suppl. Table 3:** Characteristics of out-of-hospital cardiac arrest victims, stratified by sex and time group.

|  |  | Overall | 2001-2003 | 2004-2007 | 2008-2011 | 2012-2015 | 2016-2019 | Est. 19-year<br>relative risk | 95%CI RR |
| --- | --- | --- | --- | --- | --- | --- | --- | --- | --- |
| **Female** | **Number** | 757 | 86 | 142 | 151 | 151 | 227 |  |  |
|  | **Age** | 24 (17-31) | 25 (19-32) | 25 (16-32) | 26 (20-32) | 23 (16-30) | 24 (17-30) | -0.75 yr | -3.22 – 1.72 |
|  | **Public location** | 190 (33%) | 4 (44%) | 16 (29%) | 42 (32%) | 49 (33%) | 79 (35%) | 1.10 | 0.66 – 1.91 |
|  | **Shock. rhythm** | 125 (18%) | 15 (19%) | 14 (11%) | 25 (18%) | 26 (18%) | 45 (21%) | 1.57 | 0.90 – 2.78 |
|  | **Bystander**<br>**witnessed** | 264 (35%) | 22 (26%) | 46 (32%) | 48 (32%) | 60 (40%) | 88 (39%) | 1.49 | 1.07 – 2.10 |
|  | **Bystander CPR** | 453 (60%) | 23 (27%) | 52 (37%) | 80 (53%) | 111 (74%) | 187 (82%) |  |  |
|  | **Bystander AED** | 26 (3.4%) | <=3 | <= 3 | <= 3 | 3 (2.0%) | 20 (8.9%) | 41.2 | 7.19 – 334.00 |
|  | **30-day survival** | 164 (22%) | 3 (3.5%) | 18 (13%) | 28 (19%) | 43 (28%) | 72 (33%) | 6.07 | 3.61 – 10.40 |
| **Male** | **Number** | 1 805 | 246 | 330 | 376 | 324 | 529 |  |  |
|  | **Age** | 26 (19-31) | 27 (21-32) | 27 (19-32) | 25 (19-31) | 25 (19-31) | 24 (19-31) | -1.56yr | -3.02 to -0.10 |
|  | **Public location** | 592 (44%) | 11 (65%) | 62 (47%) | 153 (45%) | 131 (41%) | 235 (45%) | 0.91 | 0.70 – 1.20 |
|  | **Shock. rhythm** | 314 (19%) | 50 (22%) | 48 (16%) | 63 (19%) | 59 (19%) | 94 (19%) | 1.02 | 0.73 – 1.43 |
|  | **Bystander**<br>**witnessed** | 621 (35%) | 71 (29%) | 99 (30%) | 128 (34%) | 117 (36%) | 206 (39%) | 1.46 | 1.17 – 1.81 |
|  | **Bystander CPR** | 1 016<br>(56%) | 75 (30%) | 120 (36%) | 198 (53%) | 213 (66%) | 410 (78%) |  |  |
|  | **Bystander AED** | 56 (3.1%) | <= 5 | <= 5 | 5 (1.3%) | 8 (2.5%) | 37 (7.2%) | 15.00 | 5.26 – 47.3 |
|  | **30-day survival** | 282 (16%) | 10 (4.1%) | 31 (9.4%) | 44 (12%) | 61 (19%) | 136 (26%) | 6.11 | 4.09 – 19.25 |

<sup>1</sup>Median (IQR); n (%)
AED: automated external defibrillators, CPR: cardiopulmonary resuscitation

